# Prognostic biomarkers of intracerebral hemorrhage identified using targeted proteomics and machine learning algorithms

**DOI:** 10.1101/2023.12.22.23300465

**Authors:** Shubham Misra, Yuki Kawamura, Praveen Singh, Shantanu Sengupta, Manabesh Nath, Zuhaibur Rahman, Pradeep Kumar, Amit Kumar, Praveen Aggarwal, Achal K. Srivastava, Awadh K. Pandit, Dheeraj Mohania, Kameshwar Prasad, Nishant K. Mishra, Deepti Vibha

## Abstract

Early prognostication of patient outcomes in intracerebral hemorrhage (ICH) is critical for patient care. We aim to investigate protein biomarkers’ role in prognosticating outcomes in ICH patients. We assessed 22 protein biomarkers using targeted proteomics in serum samples obtained from the ICH patient dataset (N=150). We defined poor outcomes as modified Rankin scale score of 3-6. We incorporated clinical variables and protein biomarkers in regression models and random forest-based machine learning algorithms to predict poor outcomes and mortality. We report Odds Ratio (OR) or Hazard Ratio (HR) with 95% Confidence Interval (CI). We used five-fold cross-validation and bootstrapping for internal validation of prediction models. We included 149 patients for 90-day and 144 patients with ICH for 180-day outcome analyses. In multivariable logistic regression, UCH-L1 (aOR 9.23; 95%CI 2.41-35.33), alpha-2-macroglobulin (5.57; 1.26-24.59), and Serpin-A11 (9.33; 1.09-79.94) were independent predictors of 90-day poor outcome; MMP-2 (6.32; 1.82-21.90) was independent predictor of 180-day poor outcome. In multivariable Cox regression models, IGFBP-3 (aHR 2.08; 1.24-3.48) predicted 90-day and MMP-9 (1.98; 1.19-3.32) predicted 180-day mortality. Using machine learning, UCH-L1 and APO-C1 predicted 90-day mortality, and UCH-L1, MMP-9, and MMP-2 predicted 180-day mortality. Overall, random forest models outperformed regression models for predicting 180-day poor outcomes (AUC 0.89), and 90-day (AUC 0.81) and 180-day mortality (AUC 0.81). Serum biomarkers independently predicted short-term poor outcomes and mortality after ICH. Further research utilizing a multiomics platform and temporal profiling is needed to explore additional biomarkers and refine predictive models for ICH prognosis.

## Introduction

Intracerebral hemorrhage (ICH) comprises 10-15% of all strokes [1], with one-month mortality of 40%, rendering it the deadliest stroke subtype [2]. It is a significant healthcare challenge worldwide, and its impact is particularly pronounced in developing nations like India. Limited healthcare resources, disparities in access to care, and unique demographic and epidemiological factors exacerbate the burden of ICH in these regions [3]. Understanding factors influencing ICH patient outcomes is crucial for optimizing clinical management, risk stratification, and resource allocation. While various clinical scores exist for predicting functional outcomes and mortality in ICH [4], such as the widely-used ICH score,^5^ their accuracy for outcomes beyond hospital discharge or 30 days remains uncertain. Hence, there is a critical need for robust prediction models that integrate new predictor variables to improve ICH prognostication [5]. Serum biomarkers have emerged as promising candidates with the potential to enhance outcome prognostication in ICH patients [6]. Integrating serum biomarkers with clinical variables in prediction models may provide additional prognostic information and guide treatment decisions.

Therefore, we undertook this study to build prognostic models using protein biomarkers to predict poor functional outcomes and mortality in ICH patients within 24 hours of symptom onset utilizing targeted proteomics, regression modeling, and machine learning approaches.

## Methods

### Study sample

We used clinical and proteomic data lodged within a prospective cohort study from a collaborative effort of the Department of Neurology, All India Institute of Medical Sciences (AIIMS), and the Institute of Genomics and Integrative Biology in New Delhi, India. The study database includes consecutive ICH patients aged ≥18 years, recruited between 04 October 2017 and 20 March 2020 within 24 hours of symptom onset. The details of this study protocol are reported in prior publications [7,8]. We obtained written informed consent from all the recruited patients or their legally authorized representatives prior to collecting blood samples and clinical history. The study was approved by the Local Institutional Ethics Committee of AIIMS, New Delhi (Ref. No. IECPG-395/28.09.2017).

### Outcomes

We defined poor outcomes as a modified Rankin Scale (mRS) score of 3-6. Our second outcome measure was mortality. The outcomes were ascertained by a researcher blinded to clinical data using telephonic interviews at 90 and 180 days post-ICH.

### Blood sample collection

Five ml of peripheral blood sample was taken in serum vacutainer tubes from ICH patients. For serum collection, it was left standing at room temperature for 30 minutes until clotted. It was then centrifuged at 3000g for 10 minutes, after which the serum was separated into cryovials. Five aliquots of each sample (100µl) were prepared and stored at −80°C until further analysis.

### Sample preparation

Ten µl of serum samples were used for protein precipitation. To 90µl of 1X Phosphate Buffer Saline (PBS), 10 µl serum was added and vortex mixed. Protein precipitation was performed using pre-chilled acetone. Briefly, to 100 µl protein extract, four times volume of pre-chilled acetone was added, vortex mixed and centrifuged at 15000 g for 10 minutes at 4℃. The supernatant was discarded, and the protein pellets were air-dried at room temperature and suspended in 0.1 M Tris-HCl with 8M urea, pH 8.5. Protein quantitation was performed using the Bradford assay.

### Reduction, Alkylation, and Trypsin Digestion

A total of 20 µg of protein from each sample was reduced with 25 mM of Dithiothreitol (DTT) for 30 minutes at 60℃, followed by alkylation using 55 mM of Iodoacetamide (IAA) at room temperature (in the dark) for 30 minutes. These samples were then subjected to trypsin digestion in an enzyme to substrate ratio of 1:10 (trypsin: protein) for 16-18 hours at 37℃. Finally, the tryptic peptides were vacuum dried in vacuum concentrator.

### Peptide selection for Multiple Reaction Monitoring (MRM)-based targeted proteomics

Peptide selection was performed using search results from ProteinPilot [9], PeptideAtlas [10] or in-silico generated peptides of proteins using Expasy PeptideCutter tool [11]. Peptides with +2 and +3 charges were considered for MRM and for each peptide, 5-6 fragment ions were used for identification (Table S1).

### Multiple Reaction Monitoring (MRM) data acquisition

Tryptic peptides obtained after digestion were desalted using reversed phase cartridges Oasis HLB cartridge (Waters, Milford, MA) according to the following procedure: wet cartridge with 1 × 1,000 μl of 100% acetonitrile, equilibrate with 1 × 1,000 μl of 0.1% formic acid, load acidified digest, washed peptides with 1 × 1,000 μl of 0.1% formic acid and elute with 1 × 1000 μl of 70% acetonitrile in 0.1% formic acid. The peptide mixture was dried using a vacuum centrifuge, and the peptides were resuspended in 0.1% formic acid at a final concentration of 1 μg/μl. A heavy labeled peptide for Apo A1 (QGLLPVLESF**K**; **K**=Lysine-13C6,15N2) protein was spiked-in the resolubilized plasma digest at a final concentration of 1 ng/μl.

The targeted MRM-MS [12] analysis of the tryptic peptides was performed on a TSQ Altis (Thermo Fisher, San Jose, CA). The instrument was equipped to an H-ESI ion source. A spray voltage of 3.5 keV was used with a heated ion transfer tube set at a temperature of 325°C. Chromatographic separations of peptides were performed on Vanquish UHPLC system (Thermo Fisher, San Jose, CA).

The 10 μl of the sample was injected and peptides were loaded on an ACQUITY UPLC BEH C18 column (130Å, 1.7 µm, 2.1 mm X 100 mm, Waters) from a cooled (4 °C) autosampler and separated with a linear gradient of water (buffer A) and acetonitrile (buffer B), containing 0.1% formic acid, at a flow rate of 300 µl/minute in 30 minutes gradient run with the buffer conditions given in Table S2.

The mass spectrometer was operated in Selected Reaction Monitoring (SRM) mode. For SRM acquisitions, the first quadrupole (Q1) and the third quadrupole (Q3) were operated at 0.7 and 0.7 unit mass resolution, respectively. A dwell time of 6.175 milliseconds (ms) was chosen, and acquisitions occurred over the whole gradient of 30 minutes. Argon was used as the collision gas at a nominal pressure of 1.5 mTorr. Optimized collision energies were used for each peptide.

### Bioinformatic and Statistical analyses

We analyzed the MRM-based targeted proteomics data using Skyline version 21.1 [13]. Peptide areas were spike-in normalized using a heavy labeled peptide for Apo A1 protein and log_2_ transformed. Peptides with <10% missing values were imputed using a random-forest-based missing value imputation method. Non-biological experimental variations were removed through batch correction.

Our study adhered to the Transparent reporting of a multivariable prediction model for individual prognosis or diagnosis (TRIPOD) guidelines [14]. We performed univariable logistic regression analysis using odds ratio (OR) and 95% confidence interval (CI) to assess poor outcomes during follow-up. Mortality rates were evaluated through Kaplan-Meier survival curves, and we constructed simple Cox proportional hazard models with hazard ratios (HR) and 95% CI. We conducted receiver operating characteristic (ROC) curve analyses and determined optimal cut-off points for each biomarker using the Youden Index (sensitivity + specificity-1).

#### Regression-based ICH outcome prediction models

We developed prediction models to determine independent predictors of poor outcomes and mortality in ICH. Variables with p-value <0.1 in the univariable analysis, along with demographic variables like age and sex, were included in a backward stepwise multiple logistic regression or multiple Cox regression analyses. Multicollinearity among predictor variables was assessed using the variance inflation factor (VIF), and predictors with a VIF value exceeding 2.5 were removed from the model. We evaluated the discrimination ability using the area under the curve (AUC) or c-statistic.

#### Machine Learning-based ICH outcome prediction models

We employed a random forest-based machine learning algorithm to identify additional predictors. Categorical data were encoded in binary form, while continuous data were standardized based on the population mean and standard deviation following log-normalization for skewed distributions. Train-test splits were executed in a 7:3 ratio, and random forest models with 1000 estimators were trained using the scikit-learn package. This process was repeated 1000 times, with each iteration involving a new random seed to choose the top 10 variables for prediction. Shapley values were computed using the SHAP package, and absolute means were utilized to evaluate variable importance.

#### Internal validation

We internally validated our prediction models using bootstrapping and 5-fold cross-validation.

We conducted statistical analyses using STATA software (Version 18) and R version 3.6.2. We conducted interaction network and enrichment analyses of significant biomarkers using Cytoscape version 3.10.0.

## Data availability

The mass spectrometry proteomics data have been deposited to the ProteomeXchange Consortium via the PRIDE [15] partner repository with the dataset identifier PXD032917.

## Results

The study cohort included 150 ICH patients recruited within 24 hours of ICH onset. Loss to follow-up at 90 days was 5% (n=1), and at 180 days was 7.33% (n=6). Therefore, we included 149 patients for the 90-day and 144 for the 180-day outcome analyses.

### Poor outcome at 90-day and 180-day

Of 149 ICH patients, 110 (73.82%) had a poor outcome at 90 days, and of 144 ICH patients, 97 (67.36%) had a poor outcome at 180 days. See Table 1 for clinical variables and Table S3 for protein biomarkers significantly associated with poor outcomes in the univariable analysis (p<0.1).

**Table 1:** Baseline characteristics and univariable analyses of poor outcomes at 90-day and 180-day after intracerebral hemorrhage.

| **Characteristics** | **90-day poor**<br>**outcome (N=110)** | **90-day good**<br>**outcome (N=39)** | **p-value** | **180-day poor**<br>**outcome (N=97)** | **180-day good**<br>**outcome (N=47)** | **p-value** |
| --- | --- | --- | --- | --- | --- | --- |
| Age (years)<sup>a</sup> | 60 (50-66) | 48 (42-55) | <0.001 | 60 (50-66) | 50 (45-60) | 0.004 |
| Male<sup>b</sup> | 66 (60) | 32 (82.05) | 0.02 | 58 (59.79) | 36 (76.60) | 0.05 |
| Female<sup>b</sup> | 44 (40) | 7 (17.95) |  | 39 (40.21) | 11 (23.40) |  |
| Blood sampling time from onset<br>(hours)<sup>a</sup> | 17 (12-20.3) | 17.5 (7.5-20) | 0.57 | 17.2 (12-20.3) | 16 (8-20) | 0.33 |
| Time taken to reach hospital<br>(hours)<sup>a</sup> | 4.71 (2.5-9) | 5 (1.5-9.83) | 0.70 | 4.5 (2.5-9) | 5 (1-9.8) | 0.55 |
| Any surgical procedure<sup>b</sup> | 25 (22.73) | 1 (2.56) | 0.02 | 24 (24.74) | 2 (4.26) | 0.008 |
| Antihypertensives<sup>b</sup> | 31 (28.18) | 12 (30.77) | 0.76 | 29 (29.90) | 11 (23.40) | 0.42 |
| ACE inhibitors<sup>b</sup> | 0 | 1 (2.56) | N.E. | 0 | 1 (2.13) | N.E. |
| ARB<sup>b</sup> | 6 (5.45) | 3 (7.69) | 0.62 | 6 (6.19) | 3 (6.38) | 0.96 |
| Beta blockers<sup>b</sup> | 31 (28.18) | 13 (33.33) | 0.55 | 31 (31.96) | 13 (27.66) | 0.60 |
| Calcium channel blockers<sup>b</sup> | 43 (39.09) | 22 (56.41) | 0.06 | 38 (39.18) | 24 (51.06) | 0.18 |
| Diuretics<sup>b</sup> | 66 (60) | 16 (41.03) | 0.04 | 58 (59.79) | 24 (51.06) | 0.32 |
| Hypertension<sup>b</sup> | 69 (62.73) | 29 (74.36) | 0.19 | 65 (67.01) | 29 (61.70) | 0.53 |
| Diabetes<sup>b</sup> | 23 (20.91) | 6 (15.38) | 0.46 | 23 (23.71) | 4 (8.51) | 0.04 |
| Dyslipidemia<sup>b</sup> | 9 (8.18) | 1 (2.56) | 0.25 | 9 (9.28) | 1 (2.13) | 0.15 |
| Myocardial Infarction<sup>b</sup> | 1 (0.91) | 1 (2.56) | 0.46 | 1 (1.03) | 1 (2.13) | 0.61 |
| Atrial fibrillation<sup>b</sup> | 3 (2.73) | 0 | N.E. | 2 (2.06) | 0 | N.E. |
| Current Smoking<sup>b</sup> | 27 (24.55) | 14 (35.90) | 0.17 | 23 (23.71) | 16 (36.17) | 0.19 |
| Mild alcohol intake<sup>b</sup> | 9 (8.18) | 4 (10.26) | 0.69 | 8 (8.25) | 5 (10.64) | 0.64 |
| Moderate alcohol intake<sup>b</sup> | 4 (3.64) | 3 (7.69) | 0.31 | 2 (2.06) | 5 (10.64) | 0.04 |
| Heavy alcohol intake<sup>b</sup> | 8 (7.27) | 7 (17.95) | 0.07 | 7 (7.22) | 6 (12.77) | 0.28 |
| No Exercise<sup>b</sup> | 86 (78.18) | 31 (79.49) | 0.86 | 76 (78.35) | 37 (78.72) | 0.96 |
| Low Education<sup>b</sup> | 88 (80) | 26 (66.67) | 0.09 | 79 (81.44) | 32 (68.09) | 0.08 |
| Low socio-economic status<sup>b</sup> | 39 (35.45) | 13 (33.33) | 0.81 | 32 (32.99) | 17 (36.17) | 0.71 |
| Obesity<sup>b</sup> | 12 (10.91) | 6 (15.38) | 0.46 | 11 (11.34) | 6 (12.77) | 0.80 |
| ICH Volume (ml)<sup>a</sup> | 36 (22.2-58.2) | 15 (9-25) | <0.001 | 37 (22.2-60) | 18 (9.5-35) | <0.001 |
| Intraventricular Hemorrhage<sup>b</sup> | 47 (42.73) | 7 (17.95) | 0.008 | 45 (46.39) | 9 (19.15) | 0.002 |
| Deep ICH<sup>b</sup> | 87 (79.09) | 36 (92.31) | 0.07 | 79 (81.44) | 39 (82.98) | 0.82 |
| Lobar ICH<sup>b</sup> | 16 (14.55) | 2 (5.13) | 0.14 | 13 (13.40) | 5 (10.64) | 0.64 |
| SBP (mmHg)<sup>a</sup> | 177.5 (154-196) | 170 (150-194) | 0.72 | 180 (154-196) | 170 (152-192) | 0.13 |
| DBP (mmHg)<sup>a</sup> | 100 (88-110) | 100 (90-113) | 0.44 | 100 (88-112) | 100 (90-110) | 0.67 |
| NIHSS score at admission<sup>a</sup> | 23 (15-28) | 10 (8-14) | <0.001 | 23 (18-28) | 10 (7-15) | <0.001 |
| GCS score at admission<sup>a</sup> | 8 (5-11) | 15 (11-15) | <0.001 | 8 (4-11) | 15 (11-15) | <0.001 |
| mRS score at discharge<sup>a</sup> | 5 (4-6) | 4 (3-4) | <0.001 | 5 (4-6) | 4 (3-4) | <0.001 |
| RBS (mg/dl)<sup>a</sup> | 155.21 (126-190) | 142 (109-168) | 0.02 | 152.14 (129-185) | 152.14 (116-176.47) | 0.09 |
| TLC (10<sup>3</sup>/μL)<sup>a</sup> | 10540 (9100-13796.29) | 9440 (7990-11022.33) | 0.01 | 11000 (9300-14100) | 9500 (7700-10910) | <0.001 |
| Hemoglobin (g/dl)<sup>a</sup> | 12.66 (11.3-14.1) | 13.6 (12.52-14.6) | 0.12 | 12.61 (11.2-14.2) | 13.54 (12.4-14.5) | 0.09 |
| Platelets (10<sup>3</sup>/μL)<sup>a</sup> | 172 (126.2-201.39) | 200 (167-254) | 0.09 | 177.67 (131-220) | 177.67 (134-243) | 0.87 |
| Blood Urea (mg%)<sup>a</sup> | 35.5 (25-45) | 27 (20-40.11) | 0.13 | 39.62 (26-46.49) | 28 (21-40.11) | 0.008 |
| Creatinine (mg%)<sup>a</sup> | 1 (0.8-1.4) | 0.93 (0.7-1.2) | 0.44 | 1.02 (0.83-1.5) | 0.9 (0.7-1.1) | 0.007 |
| Potassium (mM/L)<sup>a</sup> | 4 (3.7-4.47) | 3.84 (3.5-4.31) | 0.14 | 4 (3.6-4.4) | 3.9 (3.7-4.37) | 0.66 |
| Sodium (mM/L)<sup>a</sup> | 139.13 (136-142) | 139.2 (136-141) | 0.72 | 139.1 (136-142) | 139.5 (136-142) | 0.79 |
| Total Bilirubin (mg%)<sup>a</sup> | 0.75 (0.6-1.05) | 0.8 (0.6-1) | 0.73 | 0.75 (0.6-1.05) | 0.75 (0.6-1.04) | 0.91 |

a: Variables represented as Median (IQR), b: Variables represented as frequency (percentage).

Multivariable logistic regression identified lower UCH-L1 (adjusted OR 9.23; 95%CI 2.41-35.33), higher alpha-2-macroglobulin (aOR 5.57, 95%CI 1.26-24.59) and Serpin A11 levels (aOR 9.33; 95%CI 1.09-79.94) as independent predictors of 90-day poor outcome; whereas higher MMP-2 levels (aOR 6.32, 95%CI 1.82-21.90) independently predicted 180-day poor outcome among other clinical predictors. The models had an AUC of 0.95 at 90 days and 0.92 at 180 days (Figure 1a, b and Table 3).

**Figure 1:**
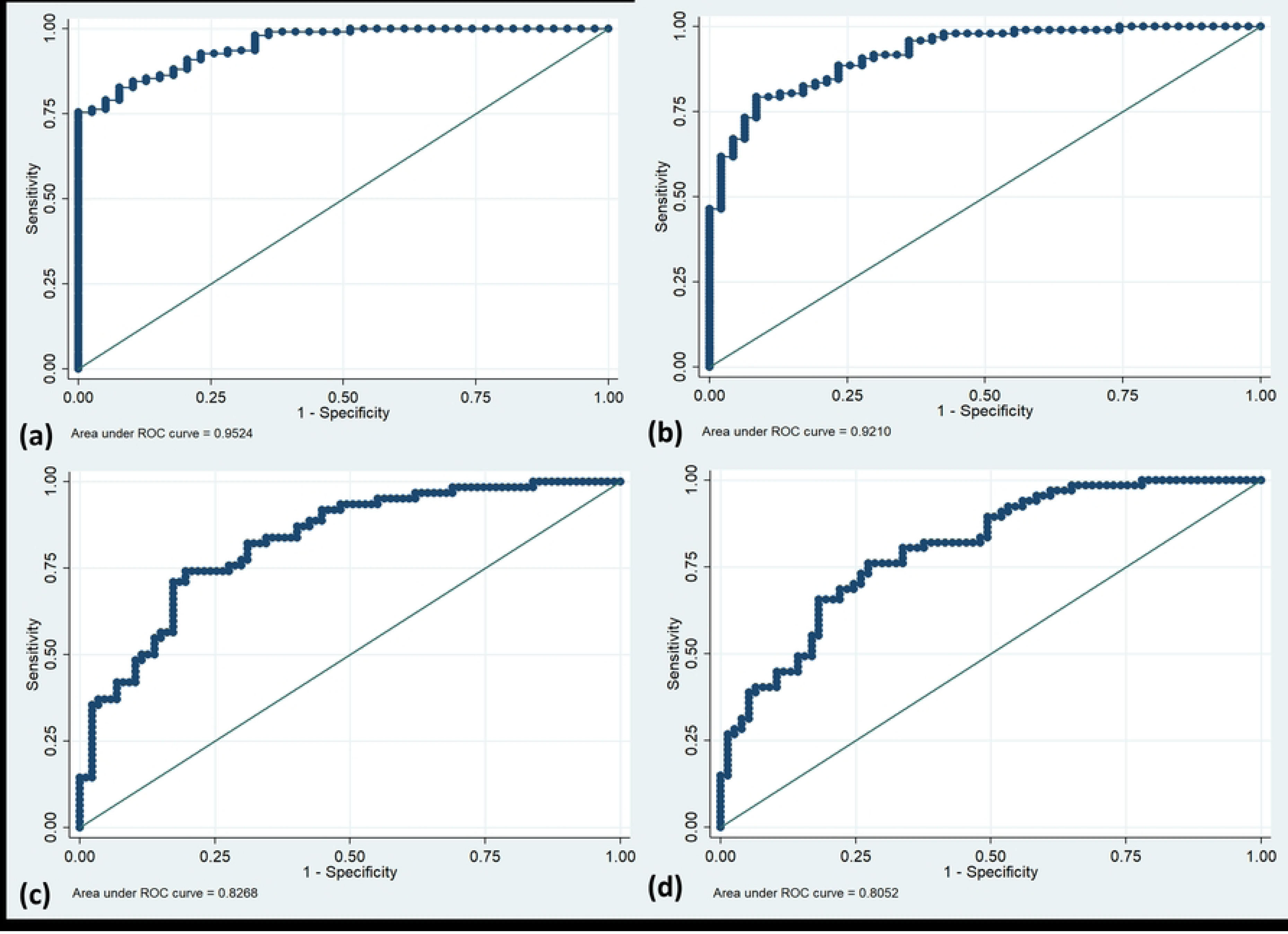
ROC curves to predict (a) 90-day poor outcome, (b) 180-day poor outcome, (c) 90-day mortality, and (d) 180-day mortality in multivariable analysis of ICH patients.

Top 10 important predictors identified by random forest algorithms for 90-day outcome included NIHSS score, ICH volume, age, UCH-L1, platelet count, RBS, alpha-2-macroglobulin, TLC, serpin A11, and haptoglobin (Figure 2a). Top 10 important predictors of 180-day poor outcome included NIHSS score, ICH volume, TLC, age, alpha-2-macroglobulin, creatinine, blood urea, UCH-L1, Serpin A11, and time taken to reach hospital (Figure 2b).

**Figure 2:**
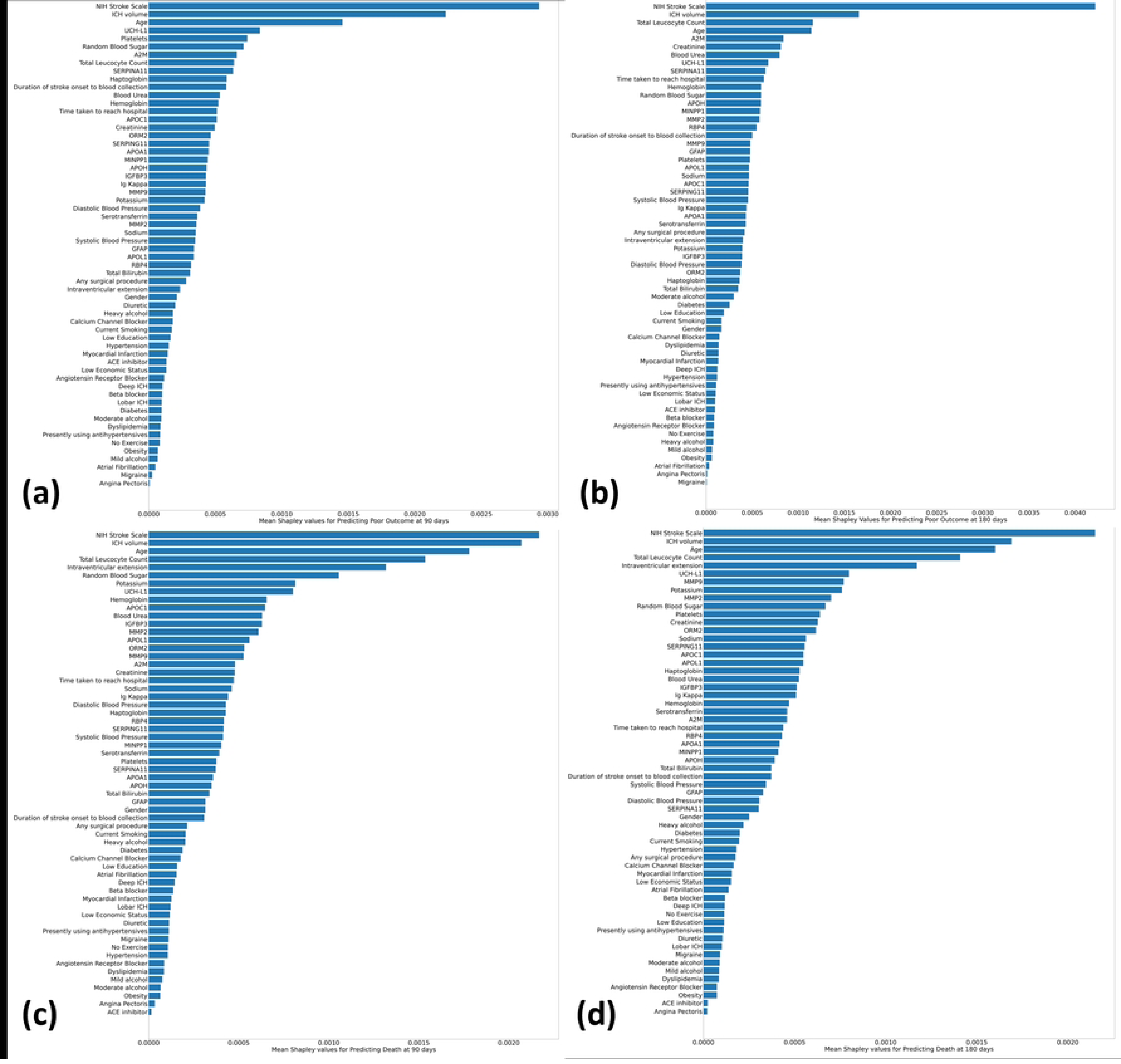
Absolute mean Shapely values calculated for random forest-based machine learning algorithms to predict (a) 90-day poor outcome, (b) 180-day poor outcome, (c) 90-day mortality, and (d) 180-day mortality in ICH.

**Figure 3:**
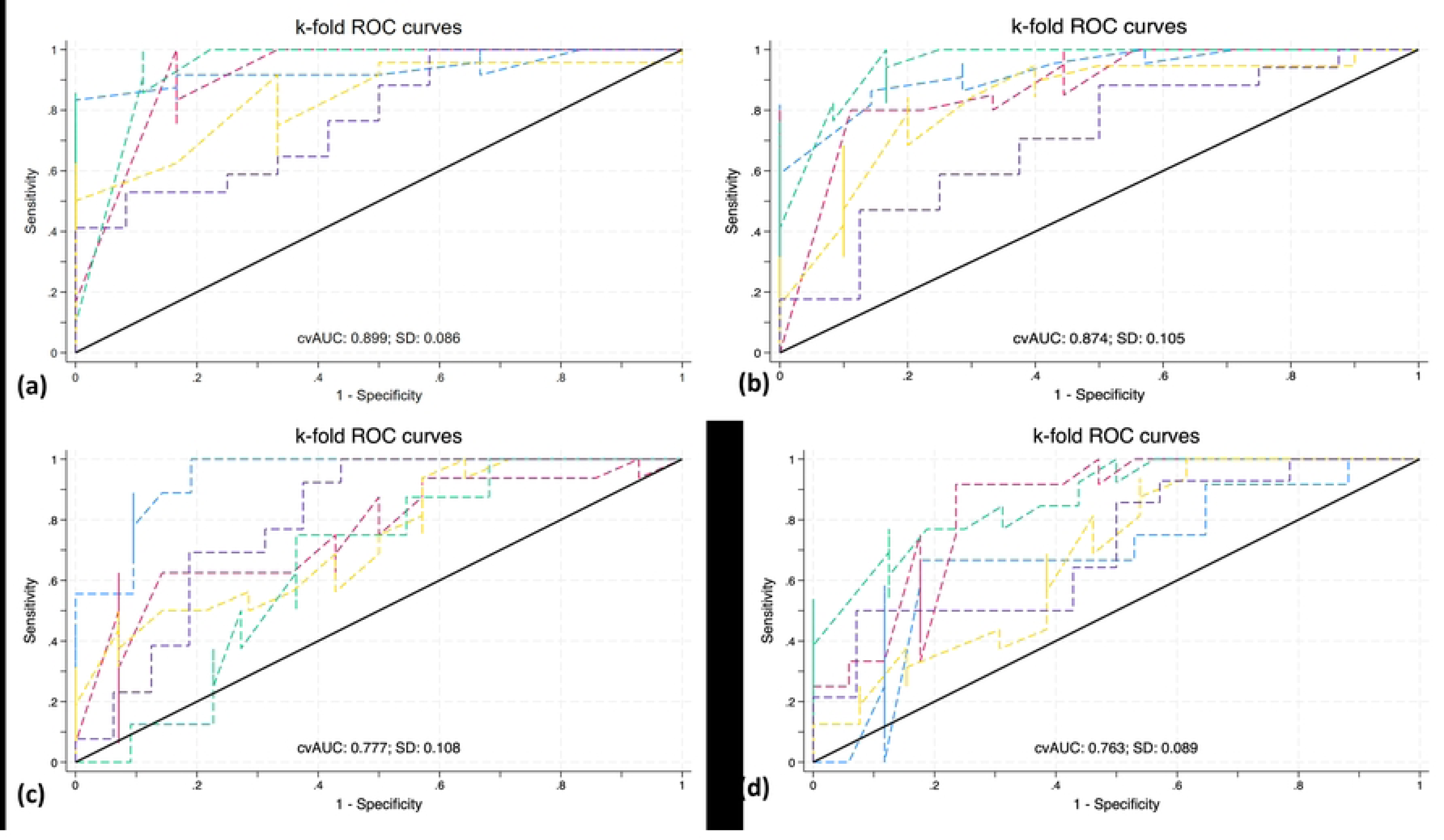
ROC curves for five-fold cross validation of multivariable regression models to predict (a) 90-day poor outcome, (b) 180-day poor outcome, (c) 90-day mortality, and (d) 180-day mortality in ICH.

**Figure 4:**
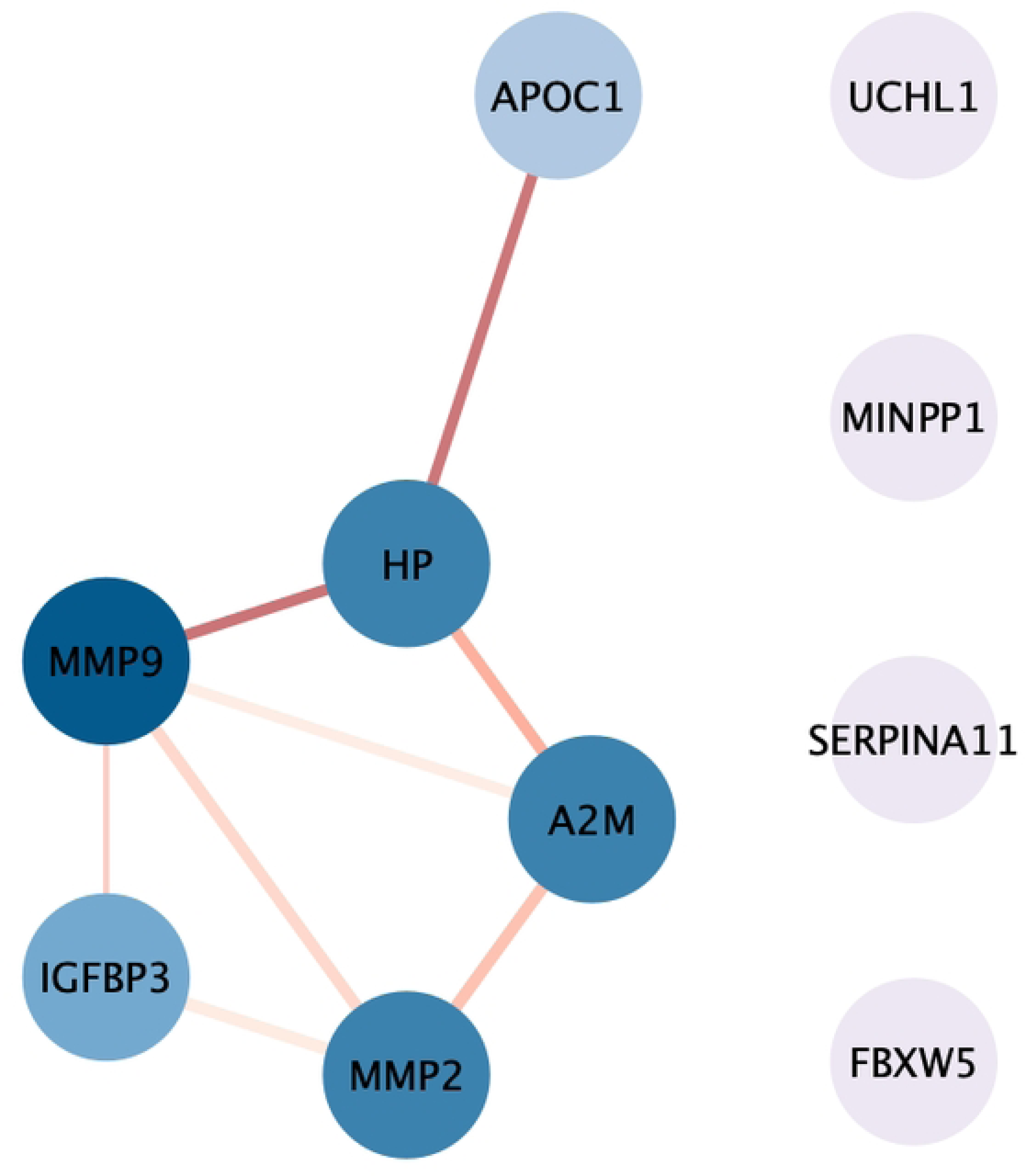
Protein-protein interaction network analysis of significant protein biomarkers that can predict poor functional outcome and mortality after ICH using the Cytoscape software. The colour of nodes and edges represents the degree of interaction and interaction score.

### Mortality at 90-day and 180-day

Mortality at 90 days was observed in 62 (41.61%) of 149 ICH patients and at 180 days in 67 (46.53%) of 144 ICH patients. See Table 2 for clinical variables and Table S4 for protein biomarkers significantly associated with mortality in the univariable analysis (p<0.1).

**Table 2:**
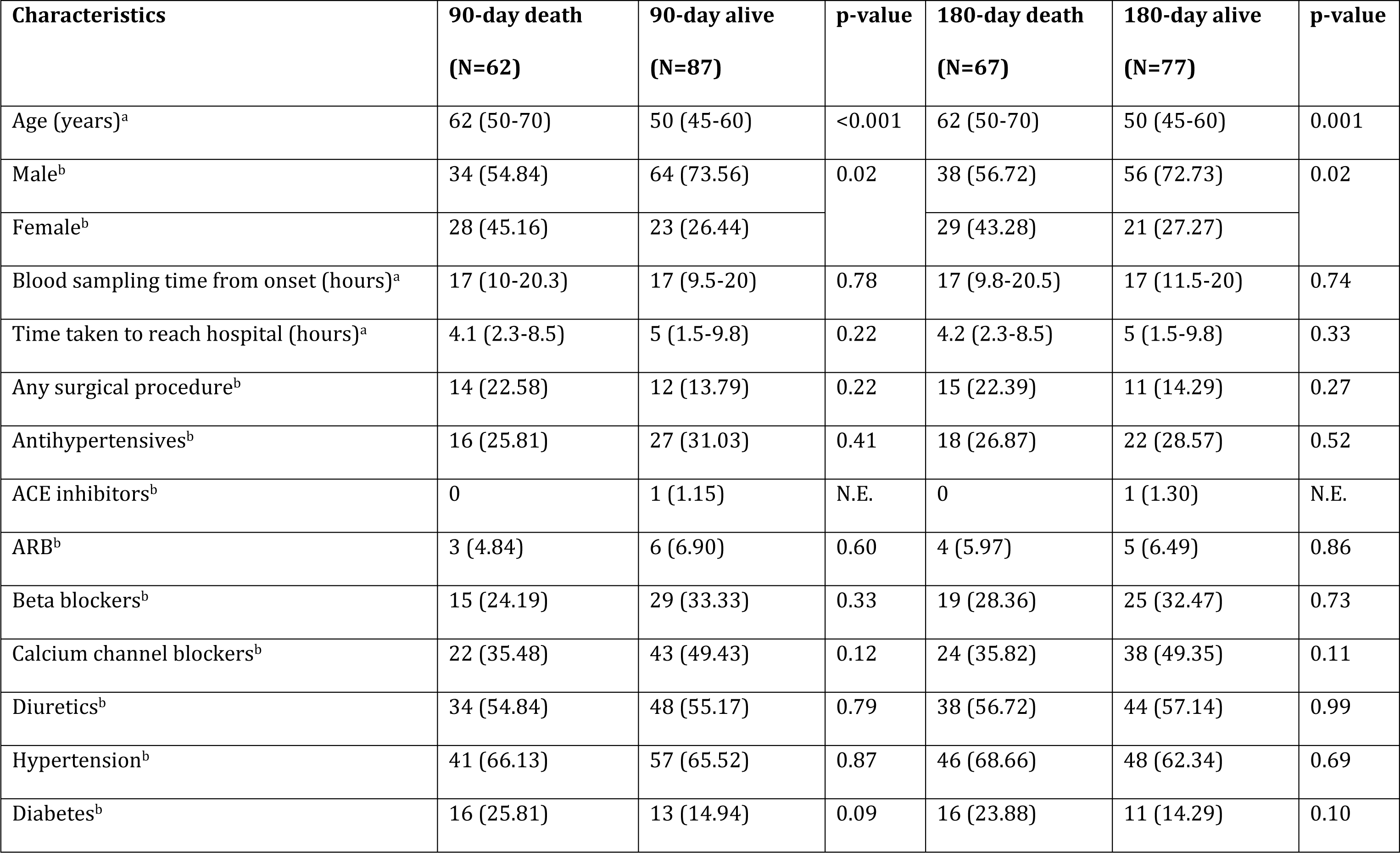
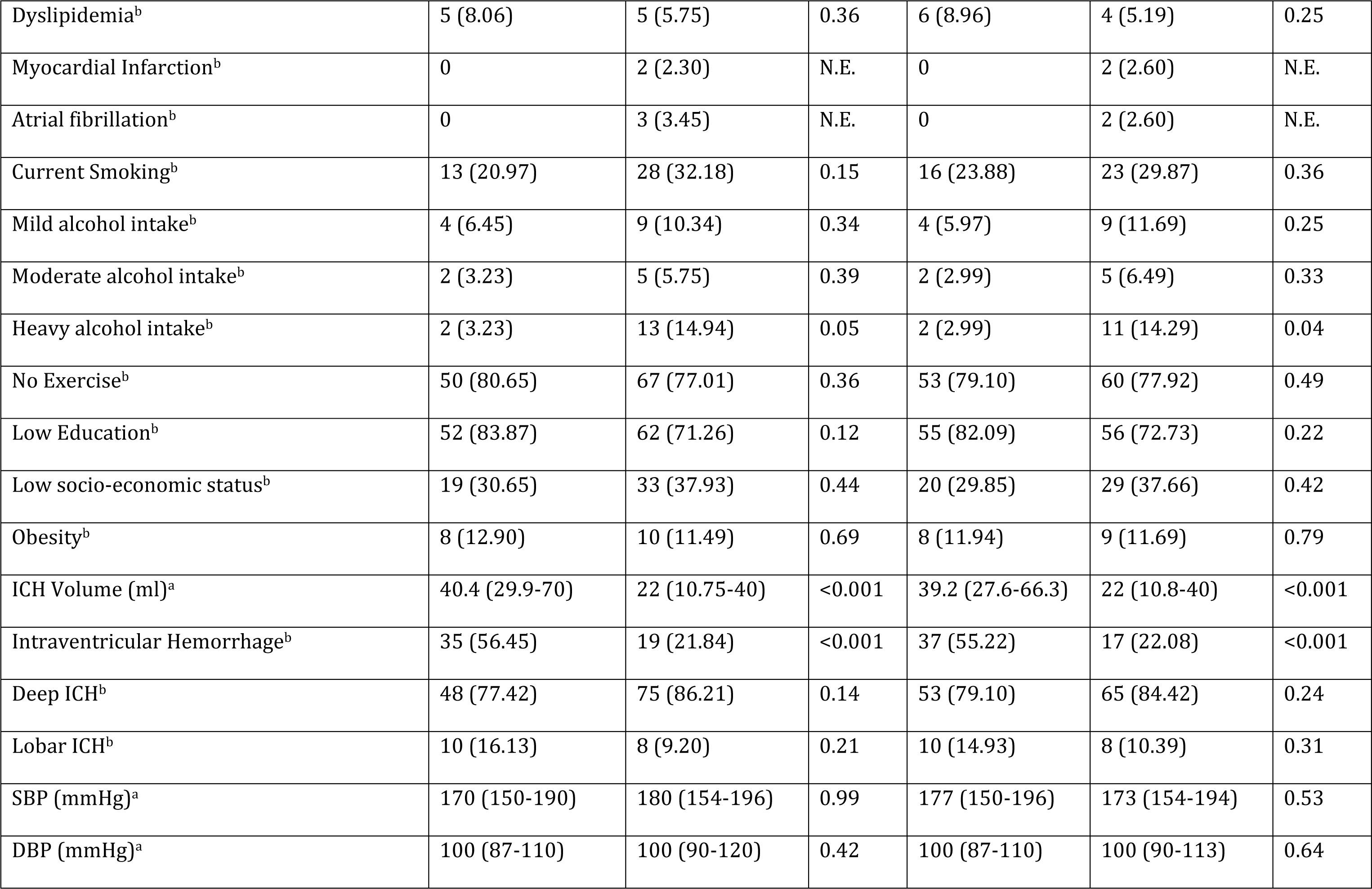
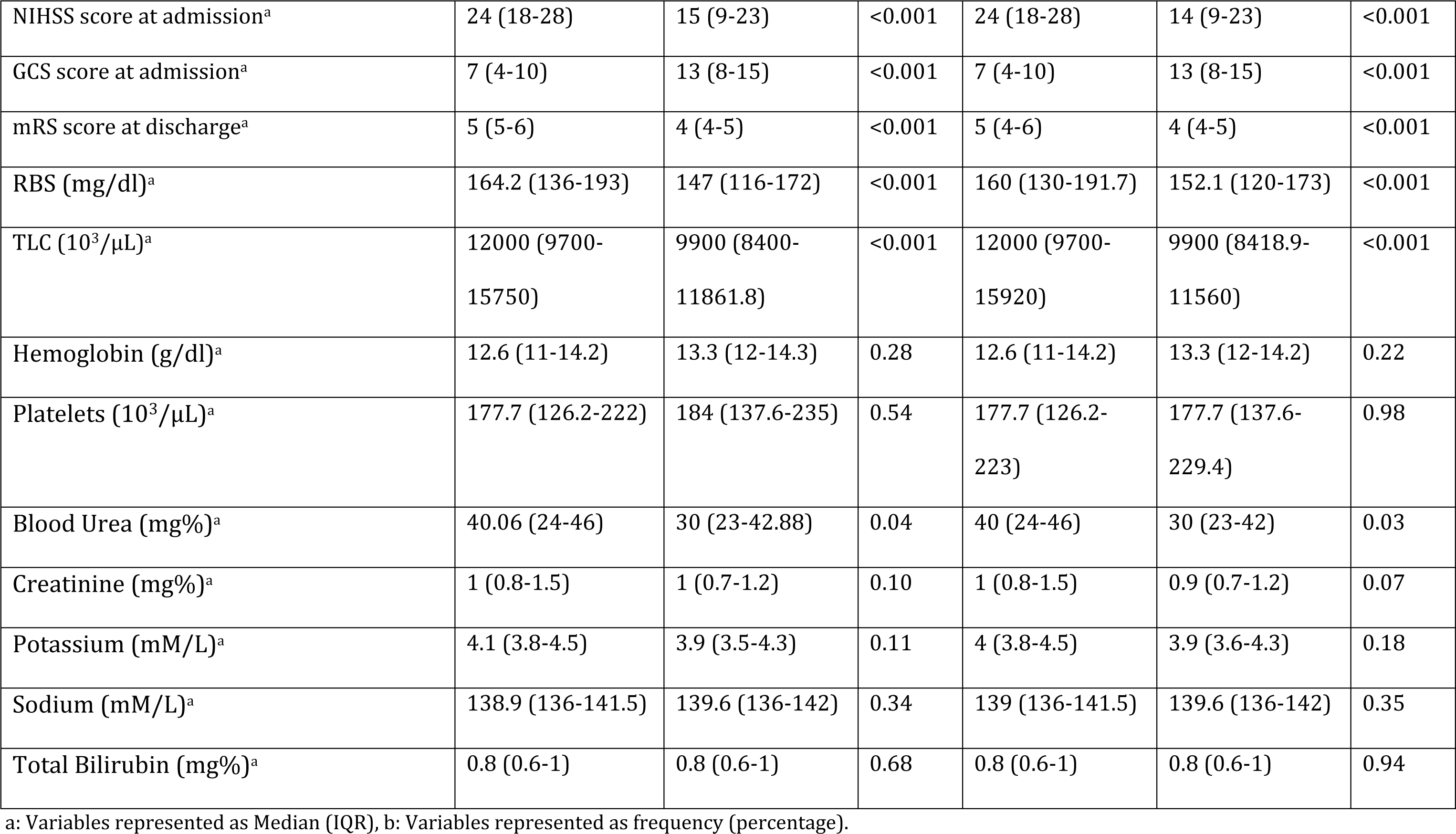
Baseline characteristics and univariable analyses of mortality at 90-day and 180-day after intracerebral hemorrhage.

| Characteristics | 90-day death<br>(N=62) | 90-day alive<br>(N=87) | p-value | 180-day death<br>(N=67) | 180-day alive<br>(N=77) | p-value |
| --- | --- | --- | --- | --- | --- | --- |
| Age (years)<sup>a</sup> | 62 (50-70) | 50 (45-60) | <0.001 | 62 (50-70) | 50 (45-60) | 0.001 |
| Male<sup>b</sup> | 34 (54.84) | 64 (73.56) | 0.02 | 38 (56.72) | 56 (72.73) | 0.02 |
| Female<sup>b</sup> | 28 (45.16) | 23 (26.44) |  | 29 (43.28) | 21 (27.27) |  |
| Blood sampling time from onset (hours)<sup>a</sup> | 17 (10-20.3) | 17 (9.5-20) | 0.78 | 17 (9.8-20.5) | 17 (11.5-20) | 0.74 |
| Time taken to reach hospital (hours)<sup>a</sup> | 4.1 (2.3-8.5) | 5 (1.5-9.8) | 0.22 | 4.2 (2.3-8.5) | 5 (1.5-9.8) | 0.33 |
| Any surgical procedure<sup>b</sup> | 14 (22.58) | 12 (13.79) | 0.22 | 15 (22.39) | 11 (14.29) | 0.27 |
| Antihypertensives<sup>b</sup> | 16 (25.81) | 27 (31.03) | 0.41 | 18 (26.87) | 22 (28.57) | 0.52 |
| ACE inhibitors<sup>b</sup> | 0 | 1 (1.15) | N.E. | 0 | 1 (1.30) | N.E. |
| ARB<sup>b</sup> | 3 (4.84) | 6 (6.90) | 0.60 | 4 (5.97) | 5 (6.49) | 0.86 |
| Beta blockers<sup>b</sup> | 15 (24.19) | 29 (33.33) | 0.33 | 19 (28.36) | 25 (32.47) | 0.73 |
| Calcium channel blockers<sup>b</sup> | 22 (35.48) | 43 (49.43) | 0.12 | 24 (35.82) | 38 (49.35) | 0.11 |
| Diuretics<sup>b</sup> | 34 (54.84) | 48 (55.17) | 0.79 | 38 (56.72) | 44 (57.14) | 0.99 |
| Hypertension<sup>b</sup> | 41 (66.13) | 57 (65.52) | 0.87 | 46 (68.66) | 48 (62.34) | 0.69 |
| Diabetes<sup>b</sup> | 16 (25.81) | 13 (14.94) | 0.09 | 16 (23.88) | 11 (14.29) | 0.10 |
| Dyslipidemia<sup>b</sup> | 5 (8.06) | 5 (5.75) | 0.36 | 6 (8.96) | 4 (5.19) | 0.25 |
| Myocardial Infarction<sup>b</sup> | 0 | 2 (2.30) | N.E. | 0 | 2 (2.60) | N.E. |
| Atrial fibrillation<sup>b</sup> | 0 | 3 (3.45) | N.E. | 0 | 2 (2.60) | N.E. |
| Current Smoking<sup>b</sup> | 13 (20.97) | 28 (32.18) | 0.15 | 16 (23.88) | 23 (29.87) | 0.36 |
| Mild alcohol intake<sup>b</sup> | 4 (6.45) | 9 (10.34) | 0.34 | 4 (5.97) | 9 (11.69) | 0.25 |
| Moderate alcohol intake<sup>b</sup> | 2 (3.23) | 5 (5.75) | 0.39 | 2 (2.99) | 5 (6.49) | 0.33 |
| Heavy alcohol intake<sup>b</sup> | 2 (3.23) | 13 (14.94) | 0.05 | 2 (2.99) | 11 (14.29) | 0.04 |
| No Exercise<sup>b</sup> | 50 (80.65) | 67 (77.01) | 0.36 | 53 (79.10) | 60 (77.92) | 0.49 |
| Low Education<sup>b</sup> | 52 (83.87) | 62 (71.26) | 0.12 | 55 (82.09) | 56 (72.73) | 0.22 |
| Low socio-economic status<sup>b</sup> | 19 (30.65) | 33 (37.93) | 0.44 | 20 (29.85) | 29 (37.66) | 0.42 |
| Obesity<sup>b</sup> | 8 (12.90) | 10 (11.49) | 0.69 | 8 (11.94) | 9 (11.69) | 0.79 |
| ICH Volume (ml)<sup>a</sup> | 40.4 (29.9-70) | 22 (10.75-40) | <0.001 | 39.2 (27.6-66.3) | 22 (10.8-40) | <0.001 |
| Intraventricular Hemorrhage<sup>b</sup> | 35 (56.45) | 19 (21.84) | <0.001 | 37 (55.22) | 17 (22.08) | <0.001 |
| Deep ICH<sup>b</sup> | 48 (77.42) | 75 (86.21) | 0.14 | 53 (79.10) | 65 (84.42) | 0.24 |
| Lobar ICH<sup>b</sup> | 10 (16.13) | 8 (9.20) | 0.21 | 10 (14.93) | 8 (10.39) | 0.31 |
| SBP (mmHg)<sup>a</sup> | 170 (150-190) | 180 (154-196) | 0.99 | 177 (150-196) | 173 (154-194) | 0.53 |
| DBP (mmHg)<sup>a</sup> | 100 (87-110) | 100 (90-120) | 0.42 | 100 (87-110) | 100 (90-113) | 0.64 |
| NIHSS score at admission<sup>a</sup> | 24 (18-28) | 15 (9-23) | <0.001 | 24 (18-28) | 14 (9-23) | <0.001 |
| GCS score at admission<sup>a</sup> | 7 (4-10) | 13 (8-15) | <0.001 | 7 (4-10) | 13 (8-15) | <0.001 |
| mRS score at discharge<sup>a</sup> | 5 (5-6) | 4 (4-5) | <0.001 | 5 (4-6) | 4 (4-5) | <0.001 |
| RBS (mg/dl)<sup>a</sup> | 164.2 (136-193) | 147 (116-172) | <0.001 | 160 (130-191.7) | 152.1 (120-173) | <0.001 |
| TLC (10<sup>3</sup>/μL)<sup>a</sup> | 12000 (9700-15750) | 9900 (8400-11861.8) | <0.001 | 12000 (9700-15920) | 9900 (8418.9-11560) | <0.001 |
| Hemoglobin (g/dl)<sup>a</sup> | 12.6 (11-14.2) | 13.3 (12-14.3) | 0.28 | 12.6 (11-14.2) | 13.3 (12-14.2) | 0.22 |
| Platelets (10<sup>3</sup>/μL)<sup>a</sup> | 177.7 (126.2-222) | 184 (137.6-235) | 0.54 | 177.7 (126.2-223) | 177.7 (137.6-229.4) | 0.98 |
| Blood Urea (mg%)<sup>a</sup> | 40.06 (24-46) | 30 (23-42.88) | 0.04 | 40 (24-46) | 30 (23-42) | 0.03 |
| Creatinine (mg%)<sup>a</sup> | 1 (0.8-1.5) | 1 (0.7-1.2) | 0.10 | 1 (0.8-1.5) | 0.9 (0.7-1.2) | 0.07 |
| Potassium (mM/L)<sup>a</sup> | 4.1 (3.8-4.5) | 3.9 (3.5-4.3) | 0.11 | 4 (3.8-4.5) | 3.9 (3.6-4.3) | 0.18 |
| Sodium (mM/L)<sup>a</sup> | 138.9 (136-141.5) | 139.6 (136-142) | 0.34 | 139 (136-141.5) | 139.6 (136-142) | 0.35 |
| Total Bilirubin (mg%)<sup>a</sup> | 0.8 (0.6-1) | 0.8 (0.6-1) | 0.68 | 0.8 (0.6-1) | 0.8 (0.6-1) | 0.94 |

a: Variables represented as Median (IQR), b: Variables represented as frequency (percentage).

**Table 3:**
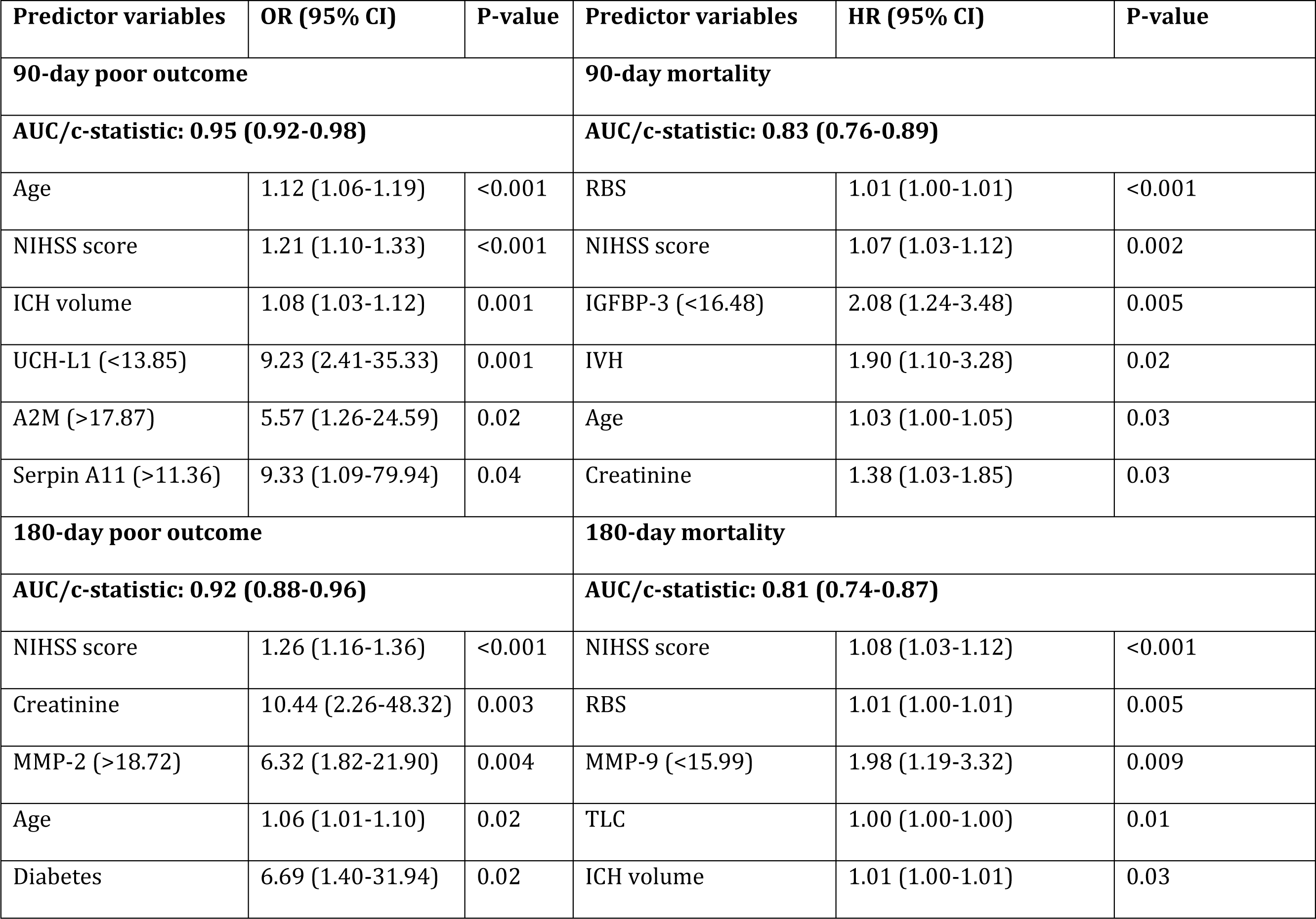
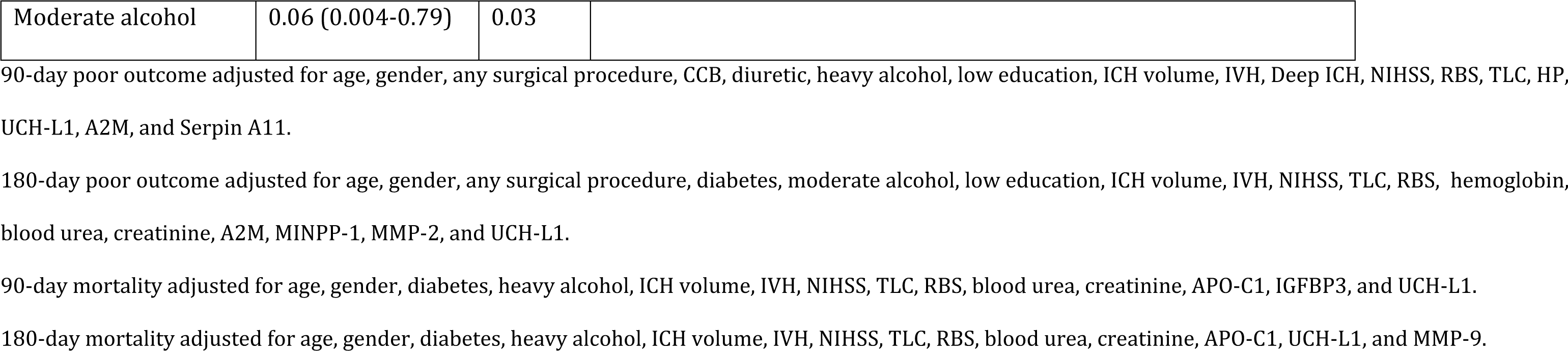
Multivariable prediction models to predict poor outcome and mortality after Intracerebral Hemorrhage.

| Predictor variables | OR (95% CI) | P-value | Predictor variables | HR (95% CI) | P-value |
| --- | --- | --- | --- | --- | --- |
| **90-day poor outcome** |  |  | **90-day mortality** |  |  |
| **AUC/c-statistic: 0.95 (0.92-0.98)** |  |  | **AUC/c-statistic: 0.83 (0.76-0.89)** |  |  |
| Age | 1.12 (1.06-1.19) | <0.001 | RBS | 1.01 (1.00-1.01) | <0.001 |
| NIHSS score | 1.21 (1.10-1.33) | <0.001 | NIHSS score | 1.07 (1.03-1.12) | 0.002 |
| ICH volume | 1.08 (1.03-1.12) | 0.001 | IGFBP-3 (<16.48) | 2.08 (1.24-3.48) | 0.005 |
| UCH-L1 (<13.85) | 9.23 (2.41-35.33) | 0.001 | IVH | 1.90 (1.10-3.28) | 0.02 |
| A2M (>17.87) | 5.57 (1.26-24.59) | 0.02 | Age | 1.03 (1.00-1.05) | 0.03 |
| Serpin A11 (>11.36) | 9.33 (1.09-79.94) | 0.04 | Creatinine | 1.38 (1.03-1.85) | 0.03 |
| **180-day poor outcome** |  |  | **180-day mortality** |  |  |
| **AUC/c-statistic: 0.92 (0.88-0.96)** |  |  | **AUC/c-statistic: 0.81 (0.74-0.87)** |  |  |
| NIHSS score | 1.26 (1.16-1.36) | <0.001 | NIHSS score | 1.08 (1.03-1.12) | <0.001 |
| Creatinine | 10.44 (2.26-48.32) | 0.003 | RBS | 1.01 (1.00-1.01) | 0.005 |
| MMP-2 (>18.72) | 6.32 (1.82-21.90) | 0.004 | MMP-9 (<15.99) | 1.98 (1.19-3.32) | 0.009 |
| Age | 1.06 (1.01-1.10) | 0.02 | TLC | 1.00 (1.00-1.00) | 0.01 |
| Diabetes | 6.69 (1.40-31.94) | 0.02 | ICH volume | 1.01 (1.00-1.01) | 0.03 |

|  |  |  |
| --- | --- | --- |
| Moderate alcohol | 0.06 (0.004-0.79) | 0.03 |

90-day poor outcome adjusted for age, gender, any surgical procedure, CCB, diuretic, heavy alcohol, low education, ICH volume, IVH, Deep ICH, NIHSS, RBS, TLC, HP, UCH-L1, A2M, and Serpin A11.
180-day poor outcome adjusted for age, gender, any surgical procedure, diabetes, moderate alcohol, low education, ICH volume, IVH, NIHSS, TLC, RBS, hemoglobin, blood urea, creatinine, A2M, MINPP-1, MMP-2, and UCH-L1.
90-day mortality adjusted for age, gender, diabetes, heavy alcohol, ICH volume, IVH, NIHSS, TLC, RBS, blood urea, creatinine, APO-C1, IGFBP3, and UCH-L1.
180-day mortality adjusted for age, gender, diabetes, heavy alcohol, ICH volume, IVH, NIHSS, TLC, RBS, blood urea, creatinine, APO-C1, UCH-L1, and MMP-9.

Multivariable Cox regression analysis identified lower IGFBP-3 levels (aHR 2.08; 95%CI 1.24-3.48) at 90 days and lower MMP-9 levels (aHR 1.98; 95%CI 1.19-3.32) at 180 days as independent predictors of mortality. The models had an AUC of 0.83 at 90 days and 0.81 at 180 days (Figure 1c, d and Table 3).

Top 10 predictors of 90-day mortality identified by random forest algorithms included NIHSS score, ICH volume, age, TLC, intraventricular extension, RBS, potassium, UCH-L1, hemoglobin, and APO-C1 (Figure 2c). Top 10 predictors of 180-day mortality included NIHSS score, ICH volume, age, TLC, intraventricular extension, UCH-L1, MMP-9, potassium, MMP-2, and RBS (Figure 2d).

### Internal validation of prediction models

Five-fold cross-validation and bootstrapping revealed that prediction models constructed with the random forest algorithm outperformed multivariable logistic and Cox regression in predicting poor outcomes at 180-day and mortality at both time points. However, there was a marginal difference in the mean AUC values of regression models and random forest models for predicting 90-day poor outcomes (Table 4, Figure S1).

**Table 4:** Five-fold cross validation and bootstrapping for internal validation of prediction models.

| **Outcome measures** | **Logistic regression (mean AUC of 5-fold cross validation)** | **Random Forest (mean AUC of 5-fold cross validation)** |
| --- | --- | --- |
| 90-day poor outcome | 0.90 (0.88-0.97) | 0.89 (0.81-0.91) |
| 180-day poor outcome | 0.87 (0.88-0.97) | 0.89 (0.85-0.95) |
| 90-day mortality | 0.78 (0.68-0.95) | 0.81 (0.73-0.89) |
| 180-day mortality | 0.76 (0.68-0.86) | 0.81 (0.78-0.85) |

### Validation of previous ICH prediction models

We validated previously published ICH prediction scores, including ICH score, MICH score, ICH-FOS score, and ICH-GS score. These scores had AUCs ranging from 0.80 to 0.86 for predicting poor outcomes at 90 days and from 0.76 to 0.84 at 180 days. For mortality prediction, the AUCs ranged from 0.78 to 0.81 at 90 days and from 0.76 to 0.80 at 180 days. Adding biomarkers to the prediction models in this study improved the prediction of poor outcomes compared to previous models, but no difference was noted in models predicting mortality (Table 5).

**Table 5:** Validation of previously published ICH prediction scores.

| S. No | Prediction score | 90-day poor outcome,<br>AUC (95% CI) | 180-day poor outcome,<br>AUC (95% CI) | 90-day mortality,<br>AUC (95% CI) | 180-day mortality,<br>AUC (95% CI) |
| --- | --- | --- | --- | --- | --- |
| 1. | ICH score [16] | 0.81 (0.74-0.88) | 0.78 (0.71-0.86) | 0.80 (0.73-0.87) | 0.80 (0.72-0.87) |
| 2. | MICH score [35] | 0.83 (0.77-0.90) | 0.80 (0.73-0.87) | 0.81 (0.73-0.88) | 0.80 (0.73-0.87) |
| 3. | ICH-FOS score [36] | 0.86 (0.79-0.92) | 0.84 (0.77-0.91) | 0.80 (0.73-0.87) | 0.80 (0.73-0.87) |
| 4. | ICH-GS score [37] | 0.80 (0.73-0.88) | 0.76 (0.67-0.84) | 0.78 (0.71-0.85) | 0.76 (0.68-0.84) |
| 5. | Present study | 0.90 (0.88-0.97) | 0.87 (0.88-0.97) | 0.78 (0.68-0.95) | 0.76 (0.68-0.86) |

**Abbreviations-** ICH: Intracerebral hemorrhage; MICH- modified Intracerebral hemorrhage; ICH-FOS: Intracerebral hemorrhage- Functional Outcome Score; ICH-GS: intracerebral hemorrhage Grading Scale; AUC: Area Under the Curve; CI: Confidence Interval.
The present study's AUCs include the internally validated 5-fold cross validation AUC values from logistic regression.
**ICH score:** Glasgow coma scale (GCS) (15-13= 0; 12-5= 1; 4-3= 2), ICH volume ( $<30 \text{ cm}^3 = 0$ ; $\geq 30 \text{ cm}^3 = 1$ ), presence of intraventricular hemorrhage (IVH) (no = 0; yes = 1), infratentorial origin (no = 0; yes = 1), age ( $<80 = 0$ ; $\geq 80 = 1$ ).
**MICH score:** GCS (15-13= 0; 12-5= 1; 4-3= 2), ICH volume ( $\leq 20 \text{ cm}^3 = 0$ ; $21-50 \text{ cm}^3 = 1$ ; $\geq 51 \text{ cm}^3 = 2$ ), presence of IVH or hydrocephalus (no = 0; yes = 1).
**ICH-FOS score:** age ( $\leq 59 = 0$ ; $60-69 = 1$ ; $70-79 = 2$ ; $\geq 80 = 4$ ), NIH Stroke Scale (NIHSS) (0-5 = 0; 6-10 = 2; 11-15 = 3; 16-20 = 4; $\geq 21 = 5$ ), GCS (15-13 = 0; 12-9 = 1; 8-3 = 2), admission glucose ( $\leq 11.0 \text{ mmol/L} = 0$ ; $\geq 11.1 \text{ mmol/L} = 1$ ), ICH volume for supratentorial origin ( $<40 \text{ cm}^3 = 0$ ; $40-70 \text{ cm}^3 = 2$ ; $>70 \text{ cm}^3 = 2$ ), ICH volume for infratentorial origin ( $<10 \text{ cm}^3 = 0$ ; $10-20 \text{ cm}^3 = 2$ ; $>20 \text{ cm}^3 = 2$ ), IVH (no = 0; yes = 1).
**ICH-GS score:** age ( $<45 = 1$ ; $45-64 = 2$ ; $\geq 65 = 3$ ), GCS (15-13 = 1, 12-9 = 2, 8-3 = 3), ICH location (supratentorial = 1; infratentorial = 2), ICH volume (for supratentorial origin: $<40 \text{ cm}^3 = 1$ ; $40-70 \text{ cm}^3 = 2$ ; $>70 \text{ cm}^3 = 3$ ; for infratentorial origin: $<10 \text{ cm}^3 = 1$ ; $10-20 \text{ cm}^3 = 2$ ; $>20 \text{ cm}^3 = 3$ ).

### Interaction network and enrichment analyses

In univariable analysis, we analyzed a protein network of 10 biomarkers linked to poor outcomes and mortality post-ICH. This network featured ten biomarkers (nodes) with eight interactions (edges), including six highly connected biomarkers. MMP-9 displayed the highest degree of interaction, connecting with four other proteins. In our network, 7 out of 8 interactions had a score exceeding 0.80, with the most robust interaction score of 0.96 between MMP-2 and IGFBP-3 (Figure S2). The significant pathways encompassed negative regulation of catalytic activity, protein metabolic processes, extracellular space, and extracellular matrix disassembly (Table S5).

## Discussion

Our analysis reaffirms the relevance of established clinical variables in predicting poor outcomes and mortality in ICH, including age, GCS score, ICH volume, NIHSS score, and various laboratory investigations, underscoring their significance in clinical practice [16,17]. Furthermore, our analyses of 22 protein biomarkers and clinical features noted in 24 hours post-ICH revealed UCH-L1, alpha-2-macroglobulin, Serpin A11, and MMP-2 as independent predictors of poor outcome. IGFBP-3 and MMP-9 independently predicted mortality following ICH. Machine learning-based random forest models identified additional predictors, including haptoglobin for poor outcomes and UCH-L1, APO-C1, and MMP-2 for mortality prediction. Integrating protein biomarkers to clinical prediction models may enhance the precision of risk assessment and resource allocation in stroke prevention programs [18], promoting more efficient resource allocation targeting individuals most likely to develop ICH.

Our study reveals inconsistent optimal values for sensitivity and specificity in biomarker cutoffs from univariable analyses (Tables S3 and S4). This emphasizes the need to consider multiple biomarkers and their cutoff values for improved accuracy and reliability in ICH outcome prediction.

This study, conducted in a tertiary care center in India, holds particular significance due to the diverse demographic, genetic, and environmental factors unique to the Indian population. Understanding stroke in this context contributes to a more holistic understanding of the disease and its multifaceted risk factors, which is vital as India and many other developing nations face an increasing burden of stroke [19]. These findings can be pivotal in shaping stroke treatment and outcome prognostication strategies unique to India and similar resource-limited settings.

UCH-L1’s association with poor outcomes in ICH suggests its potential as an early neurological damage marker [20]. Alpha-2-macroglobulin’s role in predicting poor outcomes in ICH, previously unexplored, may relate to its involvement in protease inhibition and inflammation regulation [21]. Serpin A11’s anti-inflammatory and anti-fibrotic properties may signify a response to mitigate damage after ICH [22]. IGFBP-3’s downregulation in ICH could reflect a compromised injury response [23]. The role of APO-C1 and haptoglobin in ICH prognostication, reported for the first time in our study, requires further investigation.

MMP-2 and MMP-9, implicated in tissue remodeling and inflammation [24], highlight extracellular matrix dynamics in ICH pathogenesis [25]. Indeed, our network and enrichment analyses underscored MMP-9’s central role in ICH pathophysiology (Figure S2), particularly its involvement in extracellular matrix-related pathways influencing ICH outcomes (Table S6) [25]. MMP-9’s association with short-term mortality but not poor functional outcomes aligns with previous Indian population findings [26], suggesting potential for MMP-9 inhibition as a neuroprotective strategy in ICH [27].

Prediction models are limited by the risk of overfitting to the training data, potentially compromising their generalizability. Cross-validation and bootstrapping address this by assessing model performance across different data subsets and testing stability through resampling. We, therefore, internally validated our findings through five-fold cross-validation and bootstrapping, demonstrating that random forest models consistently outperformed regression models in predicting 180-day poor outcome and 90-day and 180-day mortality, as evidenced by higher AUC values (Table 4). This suggests that machine learning algorithms, with their ability to capture complex interactions and patterns, offer a valuable tool for enhancing prognostic accuracy in ICH [28,29].

We also validated the performance of previously published prediction scores for ICH outcomes (Table S5). Our study, demonstrated higher AUC values for poor outcome predictions compared to the existing prediction scores. This suggests that integrating novel protein biomarkers and clinical features in our model enhances the precision of risk assessment for ICH outcomes.

The choice of outcomes in this study emphasizes the importance of patient-reported outcome measures (PROMs) and quality of life in stroke research and care [30]. While many clinical trials prioritize mortality as the primary outcome, it’s essential to acknowledge that survival alone may not fully represent the patient’s overall well-being or quality of life [31]. Rankin 4-5 indicates significant disability, and for many patients, this level of impairment can be as debilitating as death itself. Rankin 3 signifies moderate disability that can significantly affect a patient’s independence and quality of life. By consolidating mRS scores 3-6, our study essentially encompasses all patients experiencing death or significant disability.

Compared to previous studies, our study has several strengths. Firstly, we utilized a targeted proteomics approach, specifically multiple reaction monitoring, for precise and sensitive measurement of protein biomarkers. Secondly, we recruited patients within the 24-hour window, a critical period for stroke management, allowing earlier biomarker assessment than prior studies [26,32]. Thirdly, our study had minimal loss to follow-up (<5%).

However, our study also has several limitations. Firstly, the small sample size warrants external validation of these biomarkers in adequately powered studies. However, we internally validated our dataset and obtained consistent AUCs across the outcome measures. Secondly, our study only included patients from a single center, limiting its generalizability. Thirdly, focusing on a 24-hour time window may not capture the full ICH progression, necessitating earlier biomarker measurements [33,34] and exploring temporal changes. Lastly, we provided relative quantification data for protein biomarkers, suggesting the need for obtaining absolute quantification values in future studies.

## Conclusion

These data reflect outcomes in developing nations underscoring the potential of serum biomarkers, in conjunction with clinical variables, to enhance outcome prediction in ICH patients. Biomarkers like UCH-L1, alpha-2-macroglobulin, Serpin A11, MMP-2, IGFBP-3, and MMP-9 showed strong associations with outcomes, improving model accuracy. With better performance of random forest-based machine learning models, proteomic data holds promise for ICH prognostication. Future research should examine temporal profiles of these biomarkers in larger cohorts and explore additional pathways using multi-omics platforms to refine predictive models for ICH prognosis.

## Acknowledgements

1. S. Misra was a DST-INSPIRE Fellow supported by Department of Science and Technology, Government of India.

## Ethics Approval

The study was approved by the local institutional ethics committee of AIIMS, New Delhi (Ref. No. IECPG-395/28.09.2017). We obtained written informed consent from all the recruited patients or their legally authorized representatives prior to collecting blood samples and clinical history.

## Author Contributions

DV conceptualized the idea of this research topic, helped design the clinical methodology, and supervised each step of execution of this study. SM primarily conducted each step of this study ranging from blood sample collection, processing, proteomic experimentation, statistical and proteomic data analysis, building prediction models, results interpretation, and manuscript writing. YK conducted the machine learning analysis. SSG supervised the proteomic experimentation and its data analysis. SM and PS conducted the proteomic experiments and data analysis. ZR contributed in conducting the proteomic experimentations in the study. MN contributed in patient sample collection and processing. AK, PK, and NKM helped in statistical data analysis. DV, PA, AKS, AKP, DM, and KP aided in patient recruitment for this study. All authors contributed to the article and approved the submitted version.

## Abbreviations

ICH: Intracerebral Hemorrhage
IVH: Intraventricular hemorrhage
OR: Odds Ratio
HR: Hazard Ratio
CI: Confidence Interval
IQR: Interquartile range
AUC: Area Under the Curve
NIHSS: National Institutes of Health Stroke Scale
GCS: Glasgow Coma Scale
mRS: modified Rankin Scale
SBP: Systolic Blood Pressure
DBP: Diastolic Blood Pressure
TLC: Total Leucocyte Count
RBS: Random Blood Sugar
A2M: Alpha-2-Macroglobulin
UCH-L1: Ubiquitin carboxyl-terminal hydrolase isozyme L1
MMP: Matrix Metalloproteinase
APO: Apolipoprotein
IGFBP-3: Insulin-like growth factor-binding protein-3

## References

1. Tsao CW, Aday AW, Almarzooq ZI, Anderson CAM, Arora P, Avery CL, et al. Heart Disease and Stroke Statistics-2023 Update: A Report From the American Heart Association. Circulation. 2023;147: e93–e621. doi:10.1161/CIR.0000000000001123

2. An SJ, Kim TJ, Yoon B-W. Epidemiology, Risk Factors, and Clinical Features of Intracerebral Hemorrhage: An Update. J Stroke. 2017;19: 3–10. doi:10.5853/jos.2016.00864

3. Pandian JD, Padma Srivastava MV, Aaron S, Ranawaka UK, Venketasubramanian N, Sebastian IA, et al. The burden, risk factors and unique etiologies of stroke in South-East Asia Region (SEAR). Lancet Reg Health Southeast Asia. 2023;17: 100290. doi:10.1016/j.lansea.2023.100290

4. Gregório T, Pipa S, Cavaleiro P, Atanásio G, Albuquerque I, Chaves PC, et al. Prognostic models for intracerebral hemorrhage: systematic review and meta-analysis. BMC Med Res Methodol. 2018;18: 145. doi:10.1186/s12874-018-0613-8

5. Wartenberg KE, Hwang DY, Haeusler KG, Muehlschlegel S, Sakowitz OW, Madžar D, et al. Gap Analysis Regarding Prognostication in Neurocritical Care: A Joint Statement from the German Neurocritical Care Society and the Neurocritical Care Society. Neurocrit Care. 2019;31: 231–244. doi:10.1007/s12028-019-00769-6

6. Troiani Z, Ascanio L, Rossitto CP, Ali M, Mohammadi N, Majidi S, et al. Prognostic Utility of Serum Biomarkers in Intracerebral Hemorrhage: A Systematic Review. Neurorehabil Neural Repair. 2021;35: 946–959. doi:10.1177/15459683211041314

7. Misra S, Singh P, Sengupta S, Kushwaha M, Rahman Z, Bhalla D, et al. Subtyping strokes using blood-based biomarkers: A proteomics approach. medRxiv; 2023. p. 2023.06.10.23291233. doi:10.1101/2023.06.10.23291233

8. Misra S, Singh P, Nath M, Bhalla D, Sengupta S, Kumar A, et al. Blood-based protein biomarkers for the diagnosis of acute stroke: A discovery-based SWATH-MS proteomic approach. Front Neurol. 2022;13: 989856. doi:10.3389/fneur.2022.989856

9. Seymour SL, Hunter CL. ProteinPilot^TM^ Software overview. : 5.

10. Desiere F, Deutsch EW, King NL, Nesvizhskii AI, Mallick P, Eng J, et al. The PeptideAtlas project. Nucleic Acids Res. 2006;34: D655-658. doi:10.1093/nar/gkj040

11. Wilkins MR, Gasteiger E, Bairoch A, Sanchez JC, Williams KL, Appel RD, et al. Protein identification and analysis tools in the ExPASy server. Methods Mol Biol. 1999;112: 531–552. doi:10.1385/1-59259-584-7:531

12. Zhang H, Liu Q, Zimmerman LJ, Ham A-JL, Slebos RJC, Rahman J, et al. Methods for peptide and protein quantitation by liquid chromatography-multiple reaction monitoring mass spectrometry. Mol Cell Proteomics. 2011;10: M110.006593. doi:10.1074/mcp.M110.006593

13. MacLean B, Tomazela DM, Shulman N, Chambers M, Finney GL, Frewen B, et al. Skyline: an open source document editor for creating and analyzing targeted proteomics experiments. Bioinformatics. 2010;26: 966–968. doi:10.1093/bioinformatics/btq054

14. Collins GS, Reitsma JB, Altman DG, Moons KGM, TRIPOD Group. Transparent reporting of a multivariable prediction model for individual prognosis or diagnosis (TRIPOD): the TRIPOD statement. The TRIPOD Group. Circulation. 2015;131: 211–219. doi:10.1161/CIRCULATIONAHA.114.014508

15. Perez-Riverol Y, Csordas A, Bai J, Bernal-Llinares M, Hewapathirana S, Kundu DJ, et al. The PRIDE database and related tools and resources in 2019: improving support for quantification data. Nucleic Acids Res. 2019;47: D442–D450. doi:10.1093/nar/gky1106

16. Hemphill JC, Bonovich DC, Besmertis L, Manley GT, Johnston SC. The ICH score: a simple, reliable grading scale for intracerebral hemorrhage. Stroke. 2001;32: 891– 897. doi:10.1161/01.str.32.4.891

17. Morotti A, Phuah C-L, Anderson CD, Jessel MJ, Schwab K, Ayres AM, et al. Leukocyte Count and Intracerebral Hemorrhage Expansion. Stroke. 2016;47: 1473–1478. doi:10.1161/STROKEAHA.116.013176

18. Mishra NK, Khadilkar SV. Stroke program for India. Ann Indian Acad Neurol. 2010;13: 28–32. doi:10.4103/0972-2327.61273

19. GBD 2016 Lifetime Risk of Stroke Collaborators, Feigin VL, Nguyen G, Cercy K, Johnson CO, Alam T, et al. Global, Regional, and Country-Specific Lifetime Risks of Stroke, 1990 and 2016. N Engl J Med. 2018;379: 2429–2437. doi:10.1056/NEJMoa1804492

20. Yu W-H, Wang W-H, Dong X-Q, Du Q, Yang D-B, Shen Y-F, et al. Prognostic significance of plasma copeptin detection compared with multiple biomarkers in intracerebral hemorrhage. Clin Chim Acta. 2014;433: 174–178. doi:10.1016/j.cca.2014.03.014

21. Vandooren J, Itoh Y. Alpha-2-Macroglobulin in Inflammation, Immunity and Infections. Front Immunol. 2021;12: 803244. doi:10.3389/fimmu.2021.803244

22. Yan B, Luo L, Liu L, Wang Z, Chen R, Wu Y, et al. Serpin family proteins as potential biomarkers and therapeutic drugs in stroke: A systematic review and meta-analysis on clinical/preclinical studies. CNS Neurosci Ther. 2023;29: 1738–1749. doi:10.1111/cns.14205

23. Siddiqui EM, Mehan S, Bhalla S, Shandilya A. Potential role of IGF-1/GLP-1 signaling activation in intracerebral hemorrhage. Curr Res Neurobiol. 2022;3: 100055. doi:10.1016/j.crneur.2022.100055

24. Misra S, Talwar P, Kumar A, Kumar P, Sagar R, Vibha D, et al. Association between matrix metalloproteinase family gene polymorphisms and risk of ischemic stroke: A systematic review and meta-analysis of 29 studies. Gene. 2018;672: 180–194. doi:10.1016/j.gene.2018.06.027

25. Li H, Ghorbani S, Ling C-C, Yong VW, Xue M. The extracellular matrix as modifier of neuroinflammation and recovery in ischemic stroke and intracerebral hemorrhage. Neurobiol Dis. 2023;186: 106282. doi:10.1016/j.nbd.2023.106282

26. Sagar R, Kumar A, Verma V, Yadav AK, Raj R, Rawat D, et al. Incremental Accuracy of Blood Biomarkers for Predicting Clinical Outcomes After Intracerebral Hemorrhage. J Stroke Cerebrovasc Dis. 2021;30: 105537. doi:10.1016/j.jstrokecerebrovasdis.2020.105537

27. Dang B, Duan X, Wang Z, He W, Chen G. A Therapeutic Target of Cerebral Hemorrhagic Stroke: Matrix Metalloproteinase-9. Curr Drug Targets. 2017;18: 1358–1366. doi:10.2174/1389450118666170427151657

28. Guo R, Zhang R, Liu R, Liu Y, Li H, Ma L, et al. Machine Learning-Based Approaches for Prediction of Patients’ Functional Outcome and Mortality after Spontaneous Intracerebral Hemorrhage. J Pers Med. 2022;12: 112. doi:10.3390/jpm12010112

29. Fernandez-Lozano C, Hervella P, Mato-Abad V, Rodríguez-Yáñez M, Suárez-Garaboa S, López-Dequidt I, et al. Random forest-based prediction of stroke outcome. Sci Rep. 2021;11: 10071. doi:10.1038/s41598-021-89434-7

30. Reeves M, Lisabeth L, Williams L, Katzan I, Kapral M, Deutsch A, et al. Patient-Reported Outcome Measures (PROMs) for Acute Stroke: Rationale, Methods and Future Directions. Stroke. 2018;49: 1549–1556. doi:10.1161/STROKEAHA.117.018912

31. Heneghan C, Goldacre B, Mahtani KR. Why clinical trial outcomes fail to translate into benefits for patients. Trials. 2017;18: 122. doi:10.1186/s13063-017-1870-2

32. Ebinger M, Ipsen N, Leonards CO, Empl L, Hanne L, Liman T, et al. Circulating insulin-like growth factor binding protein-3 predicts one-year outcome after ischemic stroke. Exp Clin Endocrinol Diabetes. 2015;123: 461–465. doi:10.1055/s-0035-1554632

33. Misra S, Kumar A, Kumar P, Yadav AK, Mohania D, Pandit AK, et al. Blood-based protein biomarkers for stroke differentiation: A systematic review. Proteomics Clin Appl. 2017;11. doi:10.1002/prca.201700007

34. Misra S, Montaner J, Ramiro L, Arora R, Talwar P, Nath M, et al. Blood biomarkers for the diagnosis and differentiation of stroke: A systematic review and meta-analysis. Int J Stroke. 2020;15: 704–721. doi:10.1177/1747493020946157

35. Cho D-Y, Chen C-C, Lee W-Y, Lee H-C, Ho L-H. A new Modified Intracerebral Hemorrhage score for treatment decisions in basal ganglia hemorrhage--a randomized trial. Crit Care Med. 2008;36: 2151–2156. doi:10.1097/CCM.0b013e318173fc99

36. Ji R, Shen H, Pan Y, Wang P, Liu G, Wang Y, et al. A novel risk score to predict 1-year functional outcome after intracerebral hemorrhage and comparison with existing scores. Crit Care. 2013;17: R275. doi:10.1186/cc13130

37. Ruiz-Sandoval JL, Chiquete E, Romero-Vargas S, Padilla-Martínez JJ, González-Cornejo S. Grading scale for prediction of outcome in primary intracerebral hemorrhages. Stroke. 2007;38: 1641–1644. doi:10.1161/STROKEAHA.106.478222

